# SARS-CoV-2-specific immunity after XBB.1.5 vaccination is not influenced by subsequent influenza vaccination in dialysis patients

**DOI:** 10.64898/2025.12.16.25342339

**Authors:** Saskia Bronder, Rebecca Urschel, Felix Reinhardt, Janine Mihm, Élora Schlienger, Tina Schmidt, Susanne Brückner, Urban Sester, Martina Sester

## Abstract

Annual immunisation against both COVID-19 and seasonal influenza is now becoming standard of care, particularly ahead of anticipated winter waves. These vaccines may be co-administered on the same day or sequentially on separate days. Data on immunogenicity and the impact of consecutive vaccinations on spike-specific humoral and cellular immunity in dialysis patients remain limited.

In this real-world observational study, SARS-CoV-2-specific immune responses were evaluated in dialysis patients receiving the monovalent XBB.1.5 vaccine followed by a quadrivalent influenza vaccine 14 days later, or either vaccine alone. Specific antibodies and T-cells were quantified and characterized using enzyme-linked immunosorbent assay and flow-cytometry.

Baseline analyses from a reference-group prior to the vaccination season showed that most patients had detectable SARS-CoV-2- and influenza-specific immunity. Both the XBB.1.5 and the influenza vaccine substantially enhanced pre-existing antigen-specific humoral and cellular responses. When comparing XBB.1.5-vaccinated patients with and without subsequent influenza-vaccination, the magnitude of XBB.1.5-induced antibody or T-cell responses did not differ. Likewise, the influenza-vaccine had no non-specific effect on SARS-CoV-2-specific immune responses. Finally, spike-specific immunity remained stable over a six-month period and persisted at levels exceeding those of unvaccinated patients assessed during the same period.

In conclusion, sequential administration of COVID-19 and influenza vaccines in dialysis patients is feasible and does not compromise the immunogenicity of either vaccine. Our data are encouraging in the context of ongoing development of additional mRNA-based vaccines that may require administration in close temporal proximity to seasonal influenza immunisation, and underscore the benefit of booster vaccination in individuals with impaired immune function.

## Introduction

Infectious diseases of the lower and upper respiratory tract can be caused by rhino-, corona-, (para)influenza and respiratory syncytial viruses^1,2^. In the northern hemisphere, respiratory infectious diseases occur seasonally in the fall and winter, and slowly decline after spring^3^. Patients with end-stage chronic kidney disease or dialysis treatment are at increased risk of respiratory viral infections with severe disease and serious outcome. To protect this vulnerable group from severe disease and hospitalization, the German standing committee for vaccine recommendations (STIKO) recommends annual influenza and COVID-19 booster vaccinations with vaccines adapted to the predominant variants against influenza and SARS-CoV-2^4^. Due to uremic immunodeficiency and multiple comorbidities^5^, patients on dialysis are known to show variably impaired immune responses against a variety of vaccinations such as herpes zoster^6^, influenza^7^ or COVID-19^8-10^. Nevertheless, we have recently shown that the bivalent BA.4/5 mRNA vaccine administered in the winter season 2022 induced IgG, neutralizing titers, and specific CD4 and CD8 T-cell levels in dialysis patients to a comparable extent as in immunocompetent individuals with even higher levels of CD4 T cells in patients with a history of SARS-CoV-2 infection^11^. In subsequent seasons, the Omicron subvariant XBB.1.5 and JN.1 dominated and led to an increased incidence of breakthrough infections, due to a strongly reduced neutralizing antibody activity toward these variants in both dialysis patients^12^ and healthy individuals^13,14^. Starting with the XBB.1.5 vaccine season, the STIKO recommended simultaneous administration with the quadrivalent influenza vaccine^15^, despite the fact that studies on co-administration of the novel mRNA vaccines with other standard protein-based vaccines were rare or yielded conflicting results. Some studies in health care workers and immunocompetent adults >60 years of age showed lower antibody levels and a lower quantitative and functional antibody response after co-administration compared to a COVID-19 booster alone^16,17^, while others found that co-administration had no significant effect on COVID-19-vaccine induced immunity^18,19^. The Commission on Hygiene and Infection Prevention of the German Society of Nephrology (DGfN) recommended sequential vaccinations for patients on dialysis (distance ≥14 days)^20^, although it was unknown, whether a COVID-19-vaccine induced immune response was influenced by a subsequent influenza vaccination. We therefore used a real-world setting and characterized baseline immunity towards SARS-CoV-2 and influenza in a reference group of patients on dialysis prior to vaccination. Moreover, we performed a detailed assessment of SARS-CoV-2- and influenza-specific immune responses in dialysis patients receiving the monovalent XBB.1.5-vaccine followed by quadrivalent influenza vaccination. Moreover, vaccine-induced SARS-CoV-2-specific immunogenicity was compared with patients who only had received monovalent XBB.1.5 or quadrivalent influenza vaccination, respectively. Finally, the stability of XBB.1.5-vaccine-induced immunity was analysed over a period of 6 months.

## Results

### Study population

Ninety patients undergoing hemodialysis (n=89) or continuous ambulatory peritoneal dialysis (n=1) participated in the study. Information on patient characteristics including primary disease that led to renal failure resulting in dialysis treatment, time on dialysis, previous kidney transplantation, immunosuppressive therapy, SARS-CoV-2-related history and differential blood counts are shown in table 1. The mean age of the patients was 69.3±13.7 years (69% male, 31% female). Most patients had a history of homologous mRNA vaccination, and between three and up to six prior immunisation events (including vaccinations and infections). A total of 70 patients (77.8%) had at least one immunisation event attributable to a previous SARS-CoV-2-infection. Seventy patients were available for baseline analysis of SARS-CoV-2- and influenza-specific humoral and cellular immunity (“reference group”). Fifty-seven patients were vaccinated and had post-vaccination immunological analyses performed (17 (29.8%) after XBB.1.5 only, 11 (19.3%) after influenza (Flu) only, and 29 (50.9%) after sequential XBB.1.5+Flu).

**Table 1.** Demographic and clinical characteristics of the study population.

| Characteristics |  | Dialysis patients<br>(n=90) |
| --- | --- | --- |
| Years of age, mean (SD) |  | 69.3 (13.7) |
| Sex, n (%)<sup>a</sup> |  |  |
|  | Female | 28 (31.1) |
|  | Male | 62 (68.9) |
| Type of dialysis, n (%) |  |  |
|  | Hemodialysis | 89 (98.9) |
|  | Peritoneal dialysis | 1 (1.1) |
| Time on dialysis (years), mean (SD) |  | 4.1 (5.1) |
| Cause of kidney failure, n (%) |  |  |
|  | Autoimmune-mediated nephropathy | 8 (8.9) |
|  | Chronic glomerulonephritis | 7 (7.8) |
|  | Secondary chronic renal disease | 52 (57.8) |
|  | Innate | 12 (13.3) |
|  | Other | 6 (6.7) |
|  | Unknown | 5 (5.6) |
| Previous kidney transplantation, n (%) |  | 5 (5.6) |
| Immunosuppressive therapy, n (%) |  | 10 (11.1) |
|  | Glucocorticoids, n (%) | 8 (8.9) |
|  | MMF, n (%) | 1 (1.1) |
|  | Prednisolone/MMF, n (%) | 1 (1.1) |
| Differential blood counts median (IQR) cells/μl | | |
|  | Leucocytes, n=63 | 6300 (2350) |
|  | Granulocytes, n=51 | 4675 (2208) |
|  | Monocytes, n=51 | 553 (301) |
|  | Lymphocytes, n=51 | 1162 (865) |
| Percentage of T cells median (IQR) % among lymphocytes |  | n=90 |
|  | CD4 T cells | 36.7 (15.2) |
|  | CD8 T cells | 18.5 (12.1) |
| **SARS-CoV-2-related characteristics before XBB.1.5-vaccination** |  |  |
| History of COVID-19 vaccination regimen, n (%) |  |  |
|  | mRNA only | 68 (75.6) |
|  | Vector/mRNA combination | 4 (4.4) |
|  | unknown | 18 (20) |
| Prior SARS-CoV-2-infection<sup>b</sup>, n (%) |  | 70 (77.8) |
| Total number of prior immunisation events<sup>c</sup>, n (%) |  |  |
|  | 1 | 1 (1.1) |
|  | 2 | 1 (1.1) |
|  | 3 | 9 (10) |
|  | 4 | 24 (26.7) |
|  | 5 | 27 (30) |
|  | 6 | 13 (14.4) |
|  | unknown | 15 (16.7) |

| Characteristics | Dialysis patients<br>(n=90) |
| --- | --- |
| Type of previous immunisation, n (%) |  |
| vaccination | 37 (41.1) |
| infection | 29 (32.2) |
| unknown | 24 (26.7) |
| Days since last known previous immunisation event, median (IQR) |  |
| vaccination | 359 (138) |
| infection | 365 (223) |

<sup>a</sup>Information on sex was based on individual self-declaration; <sup>b</sup>13 patients were assigned as previously infected based on NCAP-IgG positivity despite no known history of infection; <sup>c</sup>immunisation events including vaccination and infection. Abbreviations: COVID-19, coronavirus disease 2019; IQR, interquartile range; MMF, mycophenolate mofetil; SARS-CoV-2, severe acute respiratory syndrome coronavirus 2; SD, standard deviation.

The study design is illustrated in supplementary figure S1. Baseline SARS-CoV-2 or influenza-specific immunity prior to vaccination from the reference group was compared with characteristics of SARS-CoV-2-specific immune responses in all patients after monovalent XBB.1.5 vaccination (supplementary figure S1, n=46, part I), and of influenza-specific immunity after influenza vaccination (supplementary figure S1, n=40, part II), respectively, independent of whether patients received sequential or single administration of the individual vaccines. Comparative analysis of SARS-CoV-2-specific immunity in individuals with sequential vaccination, with XBB.1.5 vaccination only and with influenza vaccination was performed in part III (supplementary figure S1, part III). Finally, the stability of SARS-CoV-2-specific immunity was analysed up to six months after sequential XBB.1.5 and influenza vaccination (supplementary figure S1, n=20, part IV).

#### Part I: SARS-CoV-2-specific humoral and cellular immunity after monovalent XBB.1.5 vaccination

To evaluate SARS-CoV-2-specific humoral and cellular immune response after XBB.1.5 vaccination, vaccine-induced immunity was analysed at two different time points (46 patients, t1, n=30; t2, n=36) and compared to the non-vaccinated reference group (n=70, figure 1a). Spike-specific IgG antibodies were significantly higher in both post-vaccination groups compared to the reference group (p<0.0001, figure 1b). After vaccination, IgG levels were above detection limit of 35.2 BAU/ml in all patients and did not differ between the post-vaccination groups. Spike-specific T cells were characterized after stimulation with overlapping peptides spanning the parental spike protein and quantified flow-cytometrically based on co-expression of the activation marker CD69 and the cytokine IFNγ. In line with IgG antibodies, spike-specific CD4 T cells were significantly higher in both post-vaccination groups than in the reference group (p<0.0001, figure 1c). No differences between both post-vaccination groups were observed. Spike-specific CD8 T cells showed higher interindividual variability. When compared to the reference group (0.03% (IQR 0.11%), their median percentages tended to be higher in the group earlier after vaccination (0.09% (IQR 0.63%), p=0.267) and reached significantly higher levels later after vaccination (0.17% (IQR 0.68%), p=0.009, figure 1d). Differences were spike-specific, as SEB-reactive CD4 and CD8 T-cell levels did not differ between the groups (figures 1c and d).

**Figure 1.**
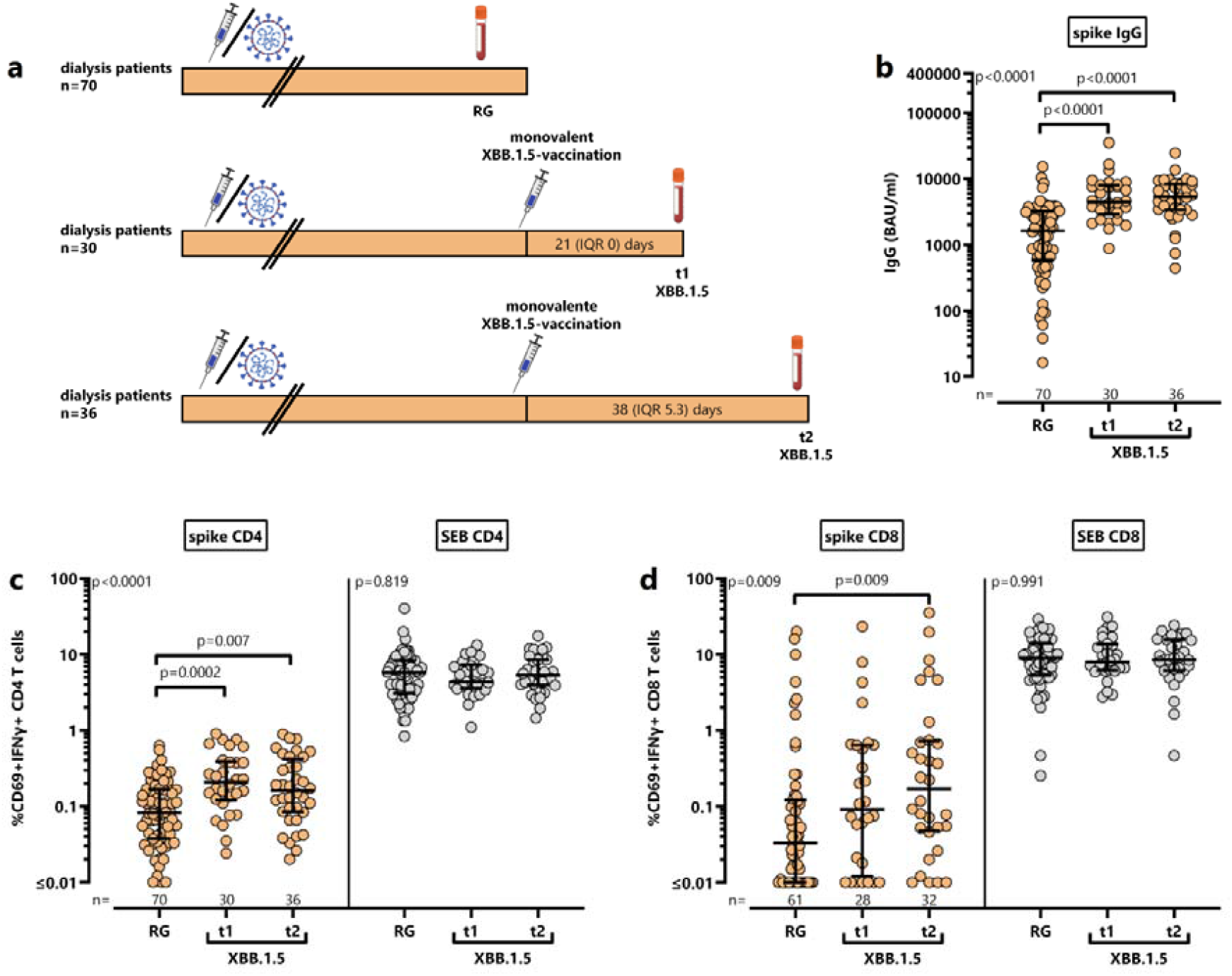
Vaccine-induced SARS-CoV-2-specific humoral and cellular immune response in dialysis patients after monovalent XBB.1.5 vaccination. **(a)** Schematic outline of the study design. Blood samples were drawn from dialysis patients (n=46) at two different time points t1 (n=30, 21 (IQR 0) days) and t2 (n=36, 38 (IQR 5.3) days) after monovalent XBB.1.5 vaccination. In addition, 70 dialysis patients without vaccination toward influenza and COVID-19 in the last three months, and without clinical evidence of SARS-CoV-2, influenza or other respiratory infections were included. Levels of spike-specific **(b)** IgG antibodies towards parental spike protein, as well as spike-specific and SEB-reactive **(c)** CD4 and **(d)** CD8 T cells were compared between the three groups. Bars represent medians with interquartile ranges. Differences were calculated using Kruskal-Wallis test followed by Dunn’s post test. Abbreviations: BAU, binding antibody unit; IFN, interferon; Ig, immunoglobulin; IQR, interquartile range; SEB, *Staphylococcus aureus* Enterotoxin B

To analyse potential differences in CD4 and CD8 T-cell levels toward spike from the parental strain and Omicron variant XBB.1.5, blood samples from a subset of patients before (n=12) and after (n=34) vaccination were also stimulated with peptide pools derived from XBB.1.5 spike. As shown in supplementary figure S2, the percentages of XBB.1.5-spike specific CD4 and CD8 T-cell levels did not differ from the respective T-cell frequencies against parental spike, indicating substantial T-cell cross reactivity between the strains.

Further phenotypical and functional characteristics of spike-specific T cells were analysed by co-expression of IFNγ, IL-2 and TNF after Boolean gating. This allowed distinction of seven subpopulations including polyfunctional cells simultaneously expressing all three cytokines, two cytokines, or one cytokine only. Spike-specific CD4 T cells were predominantly polyfunctional, followed by CD4 T cells expressing TNF alone or in combination with IFNγ or IL-2 (figure 2). Cytokine profiles of spike-specific CD4 T cells differed from those of spike-specific CD8 T cells (figure 2a). As expected, CD8 T cells produced less IL-2, and were primarily IFNγ^+^TNF^+^. The percentages of the predominant CD4 and CD8 T-cell populations were numerically lower in the early post-vaccination group than in patients later after vaccination. Furthermore, expression levels of CTLA-4 were significantly higher on spike-specific CD4 and CD8 T cells in both post-vaccination groups compared to the reference group or SEB-reactive T cells (figure 2b). In line with recent antigen encounter, median CTLA-4 expression levels of CD4 T cells were numerically higher in the group earlier after vaccination, whereas levels of CD8 T cells were rather similar in both post-vaccination groups.

**Figure 2.**
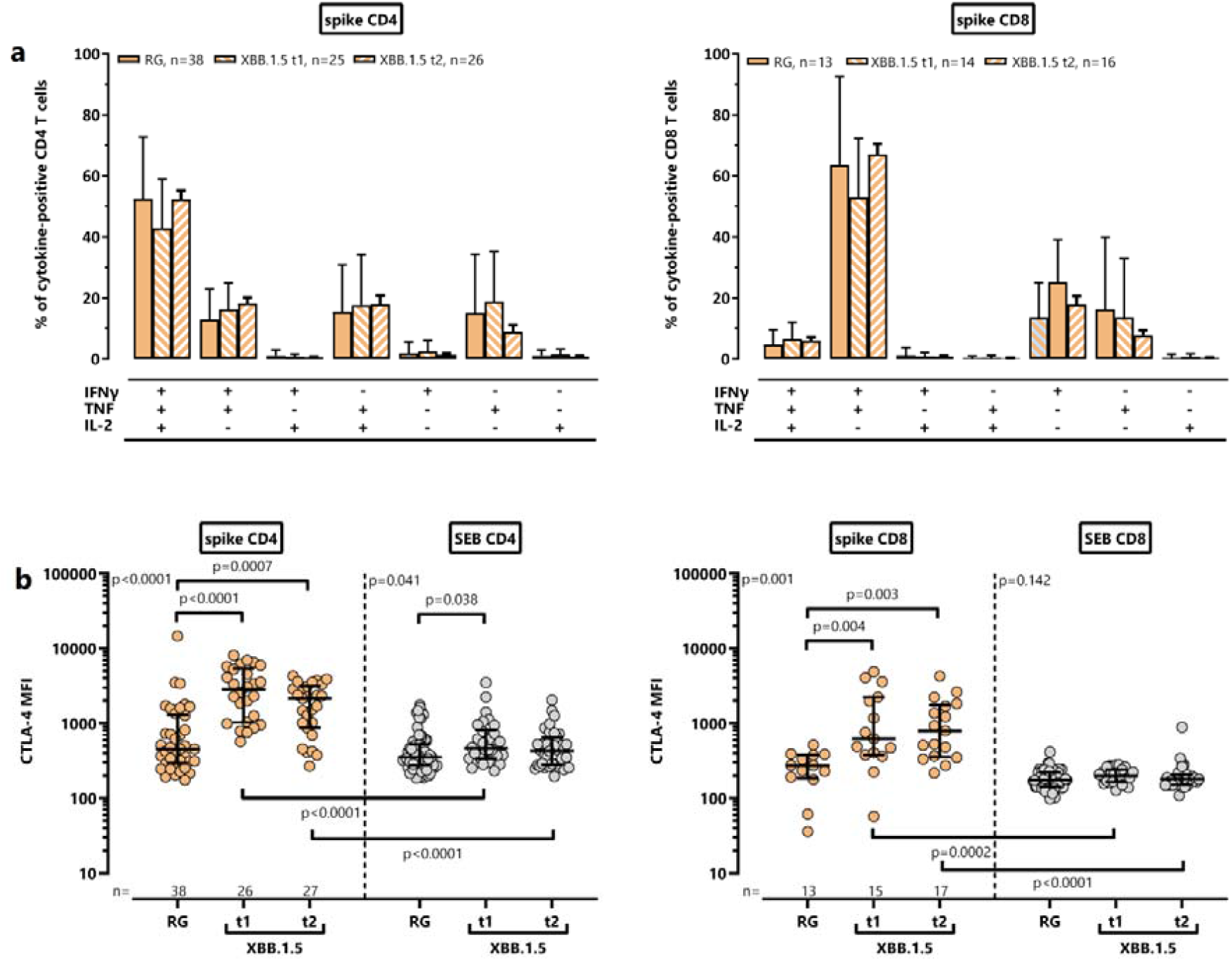
Functional and phenotypical characterization of parental spike-specific CD4 and CD8 T cells in dialysis patients after monovalent XBB.1.5. Cytokine expression profiles of **(a)** CD4 and **(b)** CD8 T cells after stimulation with parental spike protein or *Staphylococcus aureus* Enterotoxin B (SEB) were compared between both postvaccination groups t1 (n=30) and t2 (n=36) and the reference group (n=70). At the single-cell level, the cytokine-expressing T cells were differentiated into 7 subpopulations according to their expression of IFNγ, TNF and IL-2 (single, double or triple cytokine-expressing cells). Only samples of patients with at least 30 cytokine-expressing CD4 and CD8 T cells were included, respectively, to allow for robust statistical analysis. Bars represent means and standard deviations of subpopulations. Differences were determined using one-way ANOVA followed by the Bonferroni correction. Median fluorescence intensity (MFI) of CTLA-4 expression on spike-specific and SEB-reactive **(c)** CD4 and **(d)** CD8 T cells was compared. To allow robust statistical analysis, only samples with at least 20 cytokine-positive CD4 and CD8 T cells, respectively, were included. Differences between groups were analysed using Kruskal-Wallis test followed by Dunn’s post test and between stimuli using Mann–Whitney test. Abbreviations: CTLA-4, cytotoxic T-lymphocyte-associated protein 4; IFN, interferon; IL, interleukin; RG, reference group; SEB, *Staphylococcus aureus* Enterotoxin B; TNF, tumor necrosis factor

#### Part II: Influenza-specific humoral and cellular immunity after quadrivalent influenza vaccination

To evaluate influenza-specific humoral and cellular immune response after influenza vaccination, vaccine-induced immunity was analysed at two different time points (40 patients, t1, n=20; t2, n=40) and compared to the non-vaccinated reference group (n=70, figure 3a). Antibody titers directed against the two influenza A and the two influenza B components of the vaccine were examined separately. Levels of IgG antibodies toward influenza A did not differ between the groups (p=0.076), whereas median IgG antibody titers toward influenza B showed significant differences with highest levels in the late post-vaccination group p=0.002, figure 3b). Levels of IgA antibodies toward influenza A (p=0.061) and B (p=0.368) did not differ between the groups (figure 3c). Influenza-specific T cells were characterized after stimulation with the tetravalent influenza vaccine. Influenza-specific CD4 T-cell levels were significantly higher in the post-vaccination groups than in the reference group (figure 3d). In contrast, influenza-specific CD8 T-cell levels were not induced after vaccination and remained similarly low as in the baseline group (p=0.319, figure 3e). Finally, SEB-reactive CD4 or CD8 T cells did not differ between the groups.

**Figure 3.**
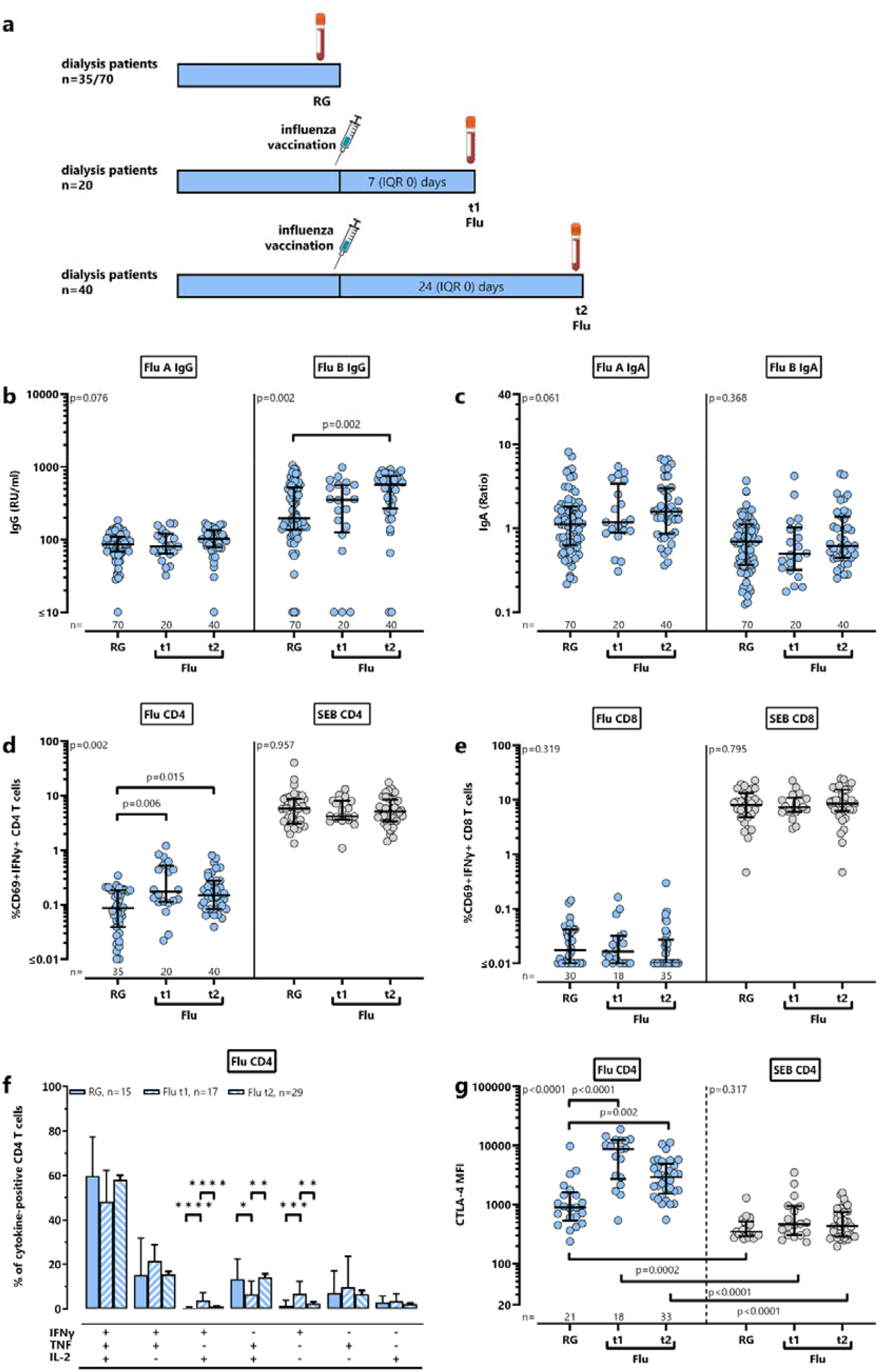
Influenza-specific humoral and cellular immunity in dialysis patients after quadrivalent influenza vaccination. **(a)** Schematic outline of the study design. Blood samples were drawn from dialysis patients (n=40) at two different time points t1 (n=20, 7 (IQR 0) days) and t2 (n=40, 24 (IQR 0) days, 11 patients with Flu only and 29 patients with XBB.1.5+Flu) after influenza vaccination (n=35 Influsplit Tetra, n=5 Efluelda). In addition, 70 dialysis patients without vaccination towards influenza and COVID-19 in the last three months, and without clinical evidence of SARS-CoV-2, influenza or other respiratory infections were included (n=35 for T-cell analyses). Levels of specific **(b)** IgG and **(c)** IgA antibodies towards influenza A and B, as well as influenza-specific and SEB-reactive **(d)** CD4 and **(e)** CD8 T cells were compared between the three groups. Bars represent medians with interquartile ranges. Differences were calculated using Kruskal-Wallis test followed by Dunn’s post test. **(f)** Cytokine expression profiles of influenza-specific CD4 T cells were compared between the three groups. At the single-cell level, the cytokine-expressing T cells were differentiated into 7 subpopulations according to their expression of IFNγ, TNF and IL-2 (single, double or triple cytokine-expressing cells). Only samples of the patients with at least 30 cytokine-expressing CD4 T cells were included to allow for robust statistical analysis. Bars represent means and standard deviations of subpopulations. Differences were determined using one-way ANOVA followed by the Bonferroni correction. **(g)** Median fluorescence intensity (MFI) of CTLA-4 expression on influenza-specific and SEB-reactive CD4 T cells was compared. To allow robust statistical analysis, only samples with at least 20 cytokine-positive CD4 T cells were included. Differences between groups were analysed using Kruskal-Wallis test followed by Dunn’s post test and between stimuli using Mann-Whitney test. Abbreviations: CTLA-4, cytotoxic T-lymphocyte-associated protein 4; Flu, influenza; IFN, interferon; Ig, immunoglobulin; IL, interleukin; IQR, interquartile range; SEB, *Staphylococcus aureus* Enterotoxin B; TNF, tumor necrosis factor

Functional analysis showed that influenza-specific CD4 T cells were mostly polyfunctional, followed by double-positive cells co-expressing either TNF/IFNγ or TNF/IL-2 (figure 3f). As with spike-specific CD4 T-cell characteristics, the cytokine profile in the immediate post-vaccination group showed a decrease in polyfunctional T cells with a concomitant shift toward higher expression of double or single IFNγ-producing populations. In the late post-vaccination group, the expression profile was comparable with that of the reference group. Again, expression of CTLA-4 was significantly higher on CD4 T cells in the post-vaccination groups than in the reference group (figure 3g). Of note, even baseline levels of CTLA-4 were higher on influenza-reactive CD4 T cells than on SEB-reactive T cells which may result from a more constant immunological challenge with influenza antigens.

#### Part III: Influence of a sequential influenza vaccination on XBB.1.5-vaccine-induced SARS-CoV-2-specific humoral and cellular immune responses

A potential influence of a sequential influenza vaccination on the XBB.1.5-induced immune response was investigated by comparing spike-specific immunity of 20 patients after sequential vaccination (“XBB.1.5+Flu”) with respective data of 17 patients receiving XBB.1.5 vaccine alone (“XBB.1.5 only”) and of 11 patients receiving influenza vaccine alone (“Flu only”, figure 4a). Despite sequential influenza vaccination after XBB.1.5, patients reached median levels of spike-specific IgG of 4442 BAU/ml (IQR 5901 BAU/ml) of similar magnitude as in patients after XBB.1.5 only (4641 BAU/ml (IQR 4407 BAU/ml)), which were significantly higher than in patients after Flu only vaccination (726 BAU/ml (IQR 1897 BAU/ml), p<0.0001, figure 4b). As with IgG, spike-specific CD4 T-cell levels were highest in patients receiving the XBB.1.5 vaccine either alone (p=0.019) or followed by the influenza vaccine (p=0.0005, figure 4c), with no significant difference between patients after XBB.1.5 vaccination alone (0.16% (IQR 0.16%)) and sequential vaccination (0.21% (IQR 0.45%)). Despite some trend toward numerically highest levels of spike-specific CD8 T cells in the XBB.1.5+Flu vaccine group, the differences between the groups did not reach statistical significance (figure 4d). Differences in CD4 T cells were spike-specific, as polyclonal T cells after SEB-stimulation were similar in the three groups.

**Figure 4.**
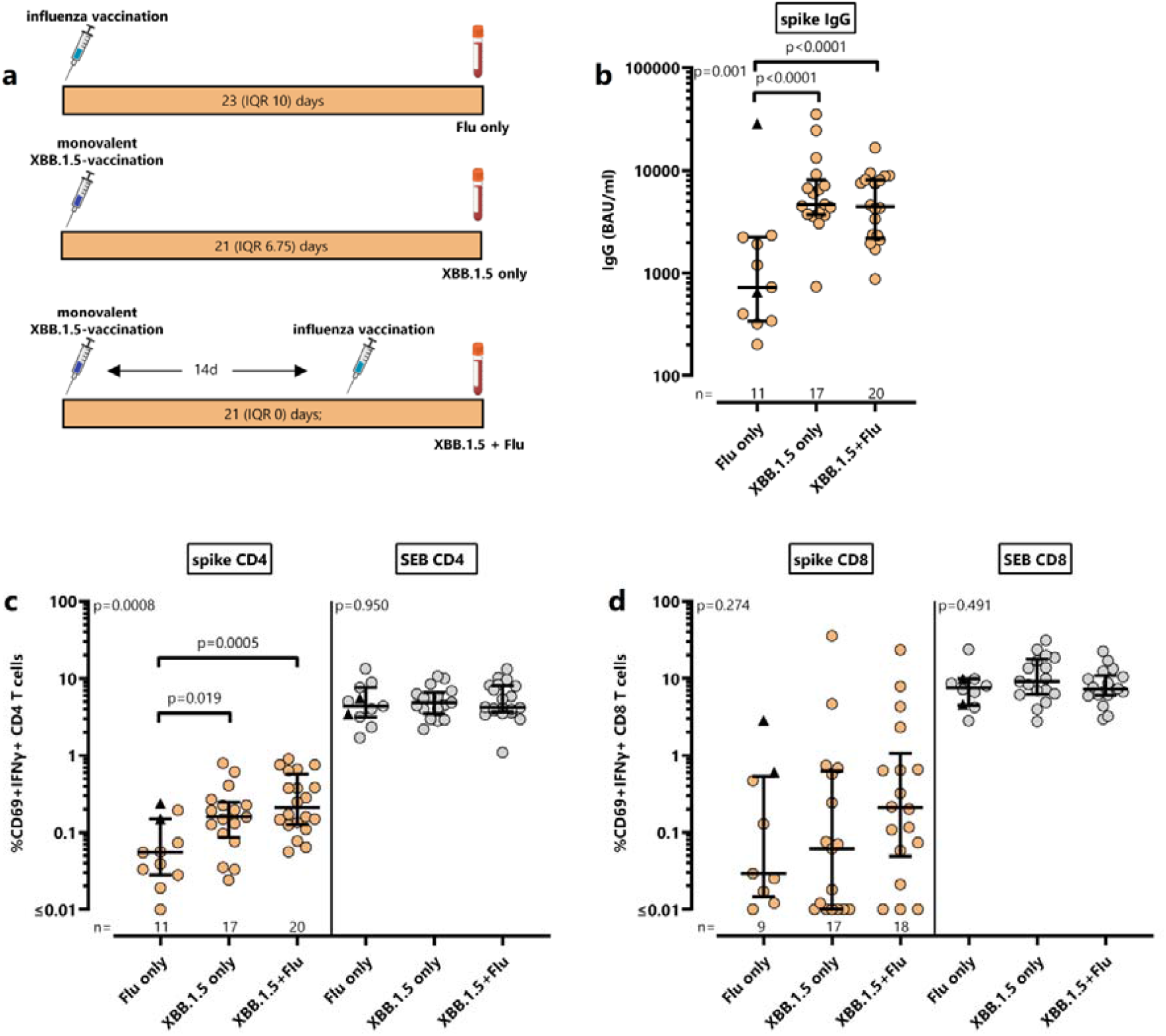
Comparison of vaccine-induced spike-specific humoral and cellular immunity in dialysis patients after sequential, monovalent XBB.1.5 alone or quadrivalent influenza vaccination alone. **(a)** Schematic outline of the study design. Blood samples were drawn from dialysis patients at a median of 23 (IQR 10) days after quadrivalent influenza vaccination (n=11), at a median of 21 (IQR 6.75) days after monovalent XBB.1.5 vaccination alone (n=17) or at a median of 21 (IQR 0) days after sequential XBB1.5+Flu vaccination (n=20, quadrivalent influenza vaccine was administered fourteen days later than XBB.1.5 vaccine). Levels of **(b)** spike-specific IgG antibodies, as well as **(c)** CD4 and **(d)** CD8 T cells after stimulation with peptides toward the parental spike protein or *Staphylococcus aureus* enterotoxin B (SEB) were compared between the three groups. Two patients, marked by a black triangle, had a previous SARS-CoV-2 infection 20 and 56 days before administration of the influenza vaccine, respectively. Bars represent medians with interquartile ranges. Differences between groups were analysed using Kruskal-Wallis test followed by Dunn’s post test. Abbreviations: BAU, binding antibody unit; Flu, influenza, IFN, interferon; Ig, immunoglobulin;

It is interesting to note that the highest spike-specific CD4 and CD8 T-cell levels in the Flu only group were observed in two patients with a history of recent SARS-CoV-2 infection (20 and 56 days before administration of the influenza vaccine, respectively, marked by a triangle, figure 4). Their median CD4 and CD8 T-cell levels were in a similar or even higher range as with those of the XBB.1.5 vaccine groups. The IgG titers of these two patients differed, with the higher titer observed in the patient with the shorter time from infection (28431.54 BAU/ml vs. 653.95 BAU/ml).

#### Part IV: Stability of spike-specific immune responses after sequential XBB.1.5+Flu vaccination

Starting from 21 (IQR 0) days after XBB.1.5 vaccination, we finally analysed the stability of the vaccine-induced spike-specific immune response two times over a 6-month time period among 20 patients who had received sequential XBB.1.5+Flu vaccination (t1/t2, n=20; t3, n=18, figure 5a).

**Figure 5.**
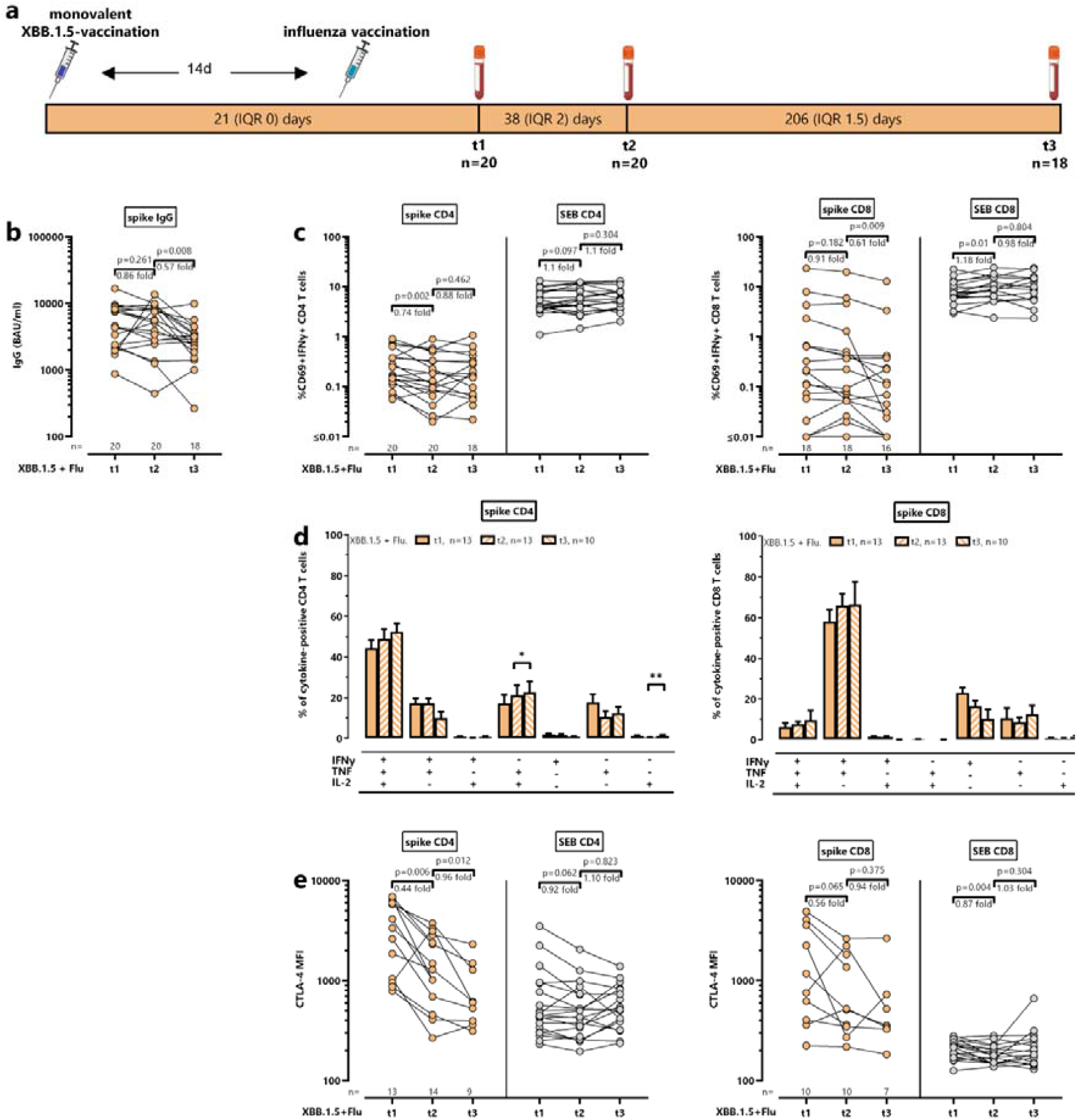
Stability of spike-specific immune responses after sequential XBB.1.5+Flu vaccination over six months. **(a)** Schematic outline of the study design. Blood samples were drawn from 20 dialysis patients receiving consecutive the monovalent XBB.1.5 vaccine and fourteen days later quadrivalent influenza vaccine (“XBB.1.5+Flu”). Starting from 21 (IQR 0) days (n=20), additional blood samples were drawn at a median of 38 (IQR 2) days (n=20) and 208 (IQR 1.5) days (n=18) after XBB.1.5 vaccination. During the 6-month period, one patient died and one patient received a kidney transplant. Levels of **(b)** spike-specific IgG antibodies, as well as **(c)** CD4 and CD8 T cells after stimulation with peptides toward the parental spike protein or *Staphylococcus aureus* enterotoxin B (SEB) were compared between the three different time points **(d)** Cytokine expression profiles of CD4 and CD8 T cells after stimulation with parental spike protein or *Staphylococcus aureus* enterotoxin B (SEB) were compared between the three different time points. At the single-cell level, the cytokine-expressing T cells were differentiated into 7 subpopulations according to their expression of IFNγ, TNF and IL-2 (single, double or triple cytokine-expressing cells). Only samples of the patients with at least 30 cytokine-expressing CD4 and CD8 T cells were included, respectively, to allow for robust statistical analysis. **(e)** Median fluorescence intensity (MFI) of CTLA-4 expression on spike-specific and SEB-reactive CD4 and CD8 T cells from patients after sequential vaccination was determined. To allow robust statistical analysis, only samples with at least 20 cytokine-positive CD4 and CD8 T cells, respectively, were included. Bars represent (b, c, e) medians with interquartile ranges or (d) means and standard deviations of subpopulations. Fold changes are indicated above the graphs and were calculated by dividing the individual levels after vaccination and levels prior to vaccination. The fold change was calculated as a ratio between post- and pre-vaccination values; to avoid division by 0 and overestimation of fold changes with low pre-vaccination levels, a value 0.03% and 0.06% was added to each percentage of CD4 and CD8 T cells prior to division, respectively. Paired analysis between time points were analysed using Wilcoxon-signed rank test. Abbreviations: CTLA-4, cytotoxic T-lymphocyte-associated protein 4; IFN, interferon; Ig, immunoglobulin; IL, interleukin; IQR, interquartile range; SEB, *Staphylococcus aureus* Enterotoxin B; TNF, tumor necrosis factor

Compared to vaccine-induced immunity after 21 days (t1), spike-specific IgG levels decreased 0.86-fold by day 38. This level further decreased 0.57-fold and reached statistical significance by 6 months after vaccination (p=0.008, figure 5b). Likewise, median percentages of spike-specific CD4 T cells showed a significant decrease after 38 days (t2, 0.74-fold, p=0.002), but remained largely stable subsequently (p=0.462, figure 5c). Spike-specific CD8 T-cell levels remained rather stable early after vaccination (t2, 0.91-fold, p=0.182), and decreased significantly by 6 months (t3, 0.61-fold, p=0.009, figure 5c). In general, no pronounced dynamic changes were observed among SEB-reactive CD4 or CD8 T-cell frequencies over time. Regarding functional and phenotypical characteristics, the overall cytokine expression profiles of spike-specific CD4 and CD8 T cells largely remained stable with some trend in increasing polyfunctionality over time (figure 5d). As expected, CTLA-4 expression on spike-specific CD4 and CD8 T cells was numerically highest early after vaccination, and decreased over the 6-month period from a median of 3345 (IQR 5070) to 604 (IQR 1005) on CD4 T cells, and from 963.5 (IQR 3312.7) to 516 (IQR 1596) on CD8 T cells after 38 days post-vaccination, with no further decrease by 6 months (0.94-fold, p=0.375, figure 5e). Despite some decrease in spike-specific immunity over a period of six months, levels of IgG antibodies and CD4 T cells remained significantly higher in patients receiving the XBB.1.5 vaccine as compared to a subgroup of 23 patients without XBB.1.5 vaccination, who were recruited in the same time period (table 2). An exception were spike-specific CD8 T cells which did not differ between the two groups (table 2).

**Table 2.** Spike-specific immune responses six months after sequential XBB.1.5+Flu vaccination compared to patients without XBB.1.5 vaccination.

| Follow-up testing | XBB.1.5+Flu<br>n=18 | No XBB.1.5 vaccination<br>n=23 | p value |
| --- | --- | --- | --- |
| Spike-specific IgG antibodies<br>(BAU/ml), median (IQR) | 2998 (1990) | 1155 (2931.9) | **0.033** |
| Spike-specific CD4 T cells<br>(% CD69<sup>+</sup>IFNγ<sup>+</sup>), median (IQR) | 0.17 (0.29) | 0.08 (0.07) | **0.021** |
| Spike-specific CD8 T cells<br>(% CD69<sup>+</sup>IFNγ<sup>+</sup>), median (IQR) | 0.09 (0.33)<br>n=16 | 0.03 (0.05) | 0.148 |

Mann-Whitney Test; during the 6-month study period, no COVID-19 infections were reported; abbreviations: BAU, binding antibody unit; IFN, Interferon; Ig, immunoglobulin; IQR, interquartile range.

## Discussion

Annual vaccination towards COVID-19 and influenza is now becoming standard of care before the seasonal waves, and the vaccines can either be co-administered on the same day or sequentially on separate days. End-stage renal disease and intermittent dialysis treatment are risk factors for severe COVID-19 and influenza disease, although knowledge of immunogenicity and the effects of consecutive vaccination series on cellular and humoral spike-specific immunity in dialysis patients is limited. We now show that both the monovalent XBB.1.5 vaccine and a quadrivalent influenza vaccine strongly induced pre-existing humoral and cellular immunity toward the respective antigens in dialysis patients. The immune responses were considerably stable over time, and we did not find any evidence that the XBB.1.5-induced immune response was adversely affected by a subsequent influenza vaccination.

Baseline analysis from a large reference group prior to the vaccine season showed that the majority of dialysis patients exhibited a SARS-CoV-2-specific immunity resulting from at least three prior antigen exposures including vaccinations and/or infections. Evidence for continued exposure with the virus was given by the fact that hybrid immunity based on a known history of infection or NCAP-specific immunity was found in more than 75% of patients, which represents a relative increase of at least 18% compared to dialysis patients recruited a year before from the same dialysis center^11^. As with spike-specific immunity, dialysis patients exhibited detectable influenza-specific antibodies and T cells at baseline, in line with regular exposure and annual vaccine recommendations^4,21^. Unlike insufficient de novo induction of immune responses in immunologically naïve patients^8-10^, we show that both the XBB.1.5 and the influenza vaccine strongly boosted pre-existing humoral and cellular immunity in dialysis patients. Regarding cellular immunity, the SARS-CoV-2-mRNA vaccine elicited both CD4 and CD8 T cells, whereas the protein-based influenza vaccine primarily induced CD4 T cells. With both vaccines, specific T cells developing at later time points were polyfunctional, whereas distinct functional and phenotypical alterations were found in the early induction phase. Early after vaccination, both vaccines led to a reduction in multifunctionality and an increase in CTLA-4 expression on the respective specific T-cell population, which reflect immunological exposure to the respective vaccine antigens. These dynamics represent typical physiological features associated with the expansion of Th_1_ cells following SARS-CoV-2^11,22-24^, and influenza vaccination^7,25,26^. Although the monovalent XBB.1.5 vaccine encodes only the spike protein of the Omicron subvariant XBB.1.5, vaccine-induced spike-specific CD4 and CD8 T cells showed pronounced cross-reactivity between XBB.1.5 and the parental SARS-CoV-2 strain. This cross-reactivity with XBB.1.5 was also found in immunocompetent individuals^27^, and with previously circulating variants such as BA.4/5 in both patients^11^ and controls^24^. Spike-specific antibodies were also strongly induced, although cross-reactivity at the level of neutralizing activity is generally less pronounced. However, other studies have reported potent XBB.1.5-vaccine induced neutralizing activity in both immunocompetent individuals^27-30^ and hemodialysis patients^12^.

Overall, we have shown that both vaccines lead to a strong immune response in dialysis patients. However, given that mRNA vaccines are still in their early development, knowledge on the translation of vaccine-antigen and immunogenicity when combined with conventional vaccines is still limited. Based on as yet poorly characterized mechanisms including trained immunity, a vaccine such as influenza^31^ or BCG^32^ may affect immunity and/or susceptibility toward infections other than the pathogen targeted by the vaccine^33^. Examples may also include herpes zoster episodes after vaccination with COVID-19 vaccines^34^ or with the inactivated shingles vaccine^35^, which both are known to have strong adjuvant activity^36-38^. A similar mechanism may also apply to immune responses upon sequential vaccinations, and may in part depend on the strength of the adjuvant effect of one vaccine that could affect induction of an immune response by the subsequent vaccine. However, results from the TACTIC study in immunocompetent individuals over 60 years of age^16^ did not show any significant differences in humoral immunity between groups receiving the COVID-19 vaccine alone, the COVID-19 vaccine followed by influenza vaccination, or the reverse vaccination sequence. These findings argue against a relevant immunological interference between an mRNA-based COVID-19 booster and a quadrivalent influenza vaccine. Likewise, our study did not provide any evidence that the quantity of XBB.1.5 vaccine-induced antibody and T-cell responses in dialysis patients were adversely affected by subsequent influenza vaccination. Furthermore, administration of the quadrivalent influenza vaccine alone did not non-specifically induce any spike-specific humoral or cellular immune response. This lack of mutual interference is further supported by the fact that phenotypical and functional changes in the early induction phase were confined to the respective T cells targeted by the vaccine and were not observed in polyclonal T-cell populations of other specificities. Despite the absence of correlates of protection, these immunological data suggest that both vaccines can be administered without compromising each other’s efficacy. These data are encouraging in light of the ongoing development of new mRNA-based vaccines toward other pathogens that may require administration in close temporal proximity to influenza vaccines.

Previous studies have described a reduced and less durable immune response following COVID-19 primary and booster immunisations in dialysis patients compared with immunocompetent individuals^39-41^. However, repeated vaccinations and natural infections contribute to the continuous maturation of immunological memory, leading to more efficient reactivation of B- and T-memory cells^42-44^. This is supported by our observation that the immune response stabilized at a level higher than non-vaccinated patients tested in the same time period. Although this prolonged stability of the immune response may result from repeated exposure to SARS-CoV-2 antigens through intermittent infections, patients did not report any COVID-19 infections during our observation period. Thus, the observed stability underscores the benefit of booster vaccinations even in individuals with impaired immune function.

A strength of our study was the first-time investigation of potential immunological interference between sequential administration of the monovalent XBB.1.5 booster and the subsequent quadrivalent influenza vaccine in dialysis patients. Moreover, patients were vaccinated during the same time period and in the same region where similar variants circulated before and after vaccination. A limitation is the restriction of antibody testing to IgG without data on neutralizing activity towards the vaccine strains. However, we have performed a detailed quantification and characterization of the cellular immune responses including stability of the booster response. A further limitation is the real-world observational setting with convenience sampling where the timing of post-vaccine sampling was primarily guided by the three routine vaccination regimens applied in the dialysis unit (i.e. either COVID-19 or influenza vaccine alone or COVID-19 followed by influenza).

In conclusion, sequential administration of the COVID-19 and influenza vaccines did not adversely affect immunogenicity of either vaccine and is a practically feasible regimen for dialysis units, where patients return on a regular basis. Unlike co-administration on the same day in two different arms, a sequential series with the use of the ipsilateral side for each vaccine is also favourable to avoid the shunt arm. We showed that the monovalent XBB.1.5 and quadrivalent influenza vaccines led to a strong induction of pre-existing baseline immunity, which is of clinical relevance regarding annual vaccine recommendations toward these respiratory pathogens. Similarly, a pronounced booster effect on specific antibodies and T cells was recently demonstrated for the newly approved mRNA- and protein-based vaccines towards the respiratory syncytial virus (RSV) in both transplant recipients^45-47^ and patients with chronic kidney disease^48^. At present, the stability of RSV-vaccine induced immunity is unknown. RSV vaccination is currently recommended once prior to the RSV infection season but may be integrated in a sequential series of respiratory seasonal vaccines. Overall, our data provide important insights for future vaccination strategies and may help refine recommendations to optimize seasonal vaccinations in immunocompromised patients.

## Methods

### Study design and subjects

In this real-world observational study, patients undergoing hemodialysis or continuous ambulatory peritoneal dialysis, and who were willing to participate were enrolled at the SHG Clinic in Völklingen, Germany. One blood sample per patient was analysed for pre-existing SARS-CoV-2- and influenza-specific humoral and cellular immunity from July to October 2023. This baseline analysis was restricted to patients without vaccination toward influenza and COVID-19 in the last three months, and without clinical evidence of SARS-CoV-2, influenza or other respiratory infections. In addition, patients receiving sequential vaccinations of Comirnaty Omicron XBB.1.5 (BioNTech/Pfizer), followed by the quadrivalent influenza vaccine (including A/Victoria/4897/2022 (H1N1)pdm09, A/Darwin/9/2021 (H3N2), B/Austria/1359417/2021, B/Phuket/3073/2013, either Influsplit Tetra (GSK) or Efluelda (Sanofi)) fourteen days later were consecutively enrolled from November 2023 to May 2024. Moreover, patients receiving the XBB.1.5 vaccine or the influenza vaccine only were recruited as control groups. Heparinized blood samples for analyses of SARS-CoV-2-and/or influenza-specific humoral and cellular immunity were collected before and at defined time points after vaccination (Supplementary figure S1). Blood samples were collected prior to a dialysis session, and vaccinations were performed after the dialysis procedure. All patients received a questionnaire for self-reporting their history of COVID-19 vaccination and infection. In addition, NCAP-IgG ELISA was performed to independently assess evidence of infection in patients without known history of COVID-19 infection. The study was performed in adherence to the declaration of Helsinki and approved by the ethics committee of the Ärztekammer des Saarlandes (reference 76/20 including amendments), and written informed consent was obtained from all patients.

### Quantitative, phenotypical and functional analysis of spike- and influenza-specific T-cell responses

To determine spike- and influenza-specific T cells, a 6h-stimulation of heparinized whole blood samples was performed as previously described^22,25^. In brief, samples were stimulated in the presence of co-stimulatory antibodies against CD28 and CD49d (clone L293 and clone 9F10, 1□μg/ml each) with overlapping peptides (each peptide 2µg/ml) spanning the parental spike or the Omicron variant XBB.1.5-spike protein (jpt Berlin, Germany), titrated amounts of the quadrivalent influenza vaccine (8µl/225µl blood; GlaxoSmithKline), 0.64% DMSO (negative control) and 2.5□μg/ml of *Staphylococcus aureus* enterotoxin B (SEB, positive control; Sigma), respectively. Brefeldin A was added at 2h. After another 4h of stimulation, cells were fixed and immunostained using anti-CD4 (clone SK3, 1:33.3), anti-CD8 (clone SK1, 1:12.5), anti-CD69 (clone L78, 1:33.3), anti-IFNγ (clone 4□S.B3, 1:100), anti-IL-2 (clone MQ1-17H12, 1:12.5), anti-TNF (clone MAb11, 1:20), and anti-CTLA-4 (clone BNI3, 1:50) and analysed using flow cytometry (BD FACS Canto II and FACSDiva software 6.1.3.). Activated CD69-positive T cells producing IFNγ identified spike- and influenza-reactive CD4 or CD8 T cells. Quantification of specific CD4 and CD8 T-cell levels was performed by subtraction of respective levels after control stimulation with detection limits of 0.03% for specific CD4 T cells and 0.06% for specific CD8 T cells as established previously^24^. T-cell functionality was characterized by analyzing co-expression of IFNγ, IL-2 and TNF as well as the cytotoxic T-lymphocyte-associated protein 4 (CTLA-4).

### Determination of SARS-CoV-2- and influenza-specific antibodies

Antibody tests were performed according to the manufacturer’s instructions (Euroimmun, Lübeck, Germany) as previously described^22,25^. SARS-CoV-2-specific IgG antibodies toward the receptor binding domain of the parental SARS-CoV-2-spike protein were quantified using the enzyme-linked immunosorbent assay (ELISA, SARS-CoV-2-QuantiVac). Antibody binding units (BAU/ml) <25.6 were scored negative, ≥25.6 and <35.2 were scored intermediate, and ≥35.2 were scored positive. Anti-SARS-CoV-2-NCP-ELISA was used to determine SARS-CoV-2-specific IgG toward the nucleocapsid (NCAP) protein. NCAP-ELISA positivity served as independent evidence for infection in patients without known COVID-19 infection history.

Influenza-specific IgA and IgG antibodies were determined using Anti-Influenza-A/B-Virus-ELISA. IgA levels represent semiquantitative values calculated by dividing the ratio between the extinction of the serum sample and the extinction of a calibration serum corresponding to the upper limit of the normal range. Ratios <0.8 were scored negative, ≥0.8 and <1.1 were scored intermediate, and ≥1.1 were scored positive. IgG levels expressed in relative units (RU/ml) were scored as negative for values <16, intermediate for values ≥16 and <22 and positive for values ≥22.

### Statistical analysis

Statistical analyses were carried out using GraphPad Prism 10.6.0 software (GraphPad, San Diego, CA, USA) using two-tailed tests. Unpaired nonparametric data between groups were compared using Mann-Whitney test or Kruskal-Wallis test followed by Dunn’s multiple comparison test. Wilcoxon matched pairs test was used to compare paired data between two groups. A p value <0.05 was considered statistically significant.

## Data Availability

All data produced in the present study are available upon reasonable request to the authors.

## Data availability

All figures and tables have associated raw data. The data that support the findings of this study are available from the corresponding author upon request.

## Acknowledgements

The authors thank the team of Department for Kidney Diseases and Hypertension at the SHG Clinic and the team of Heimdialyse Saar e.V. in Völklingen for their support in enrolling participants. The authors also thank all participants of this study. Expert technical assistance by Candida Guckelmus is acknowledged. Drawings in figures were in part generated by BioRender. Financial support was provided in part by the State chancellery of the Saarland to M.S., by the German Federal Ministry of Education and Research (COVIM 2.0, FKZ 01KX2121) to M.S., and by the Dr. Rolf M. Schwiete Stiftung (Project #2023-047) to T.S.

## Author Contributions

Sa.B., T.S., J.M. U.S., and M.S. designed the study and the experiments. Sa.B., R.U., and E.S. performed experiments. Sa.B., F.R., J.M., Su.B., and U.S. contributed to patient recruitment and clinical data acquisition. Sa.B. and M.S. performed statistical analysis. T.S., J.M., U.S. and M.S. supervised all parts of the study. Sa.B. and M.S. wrote the manuscript, and all other authors provided revisions for important intellectual content. All authors agree on accountability for all aspects of the work in ensuring that questions related to the accuracy or integrity of any part of the work are appropriately investigated and resolved. All authors approved the final version of the manuscript.

## Supplement

**Supplementary figure S1.**
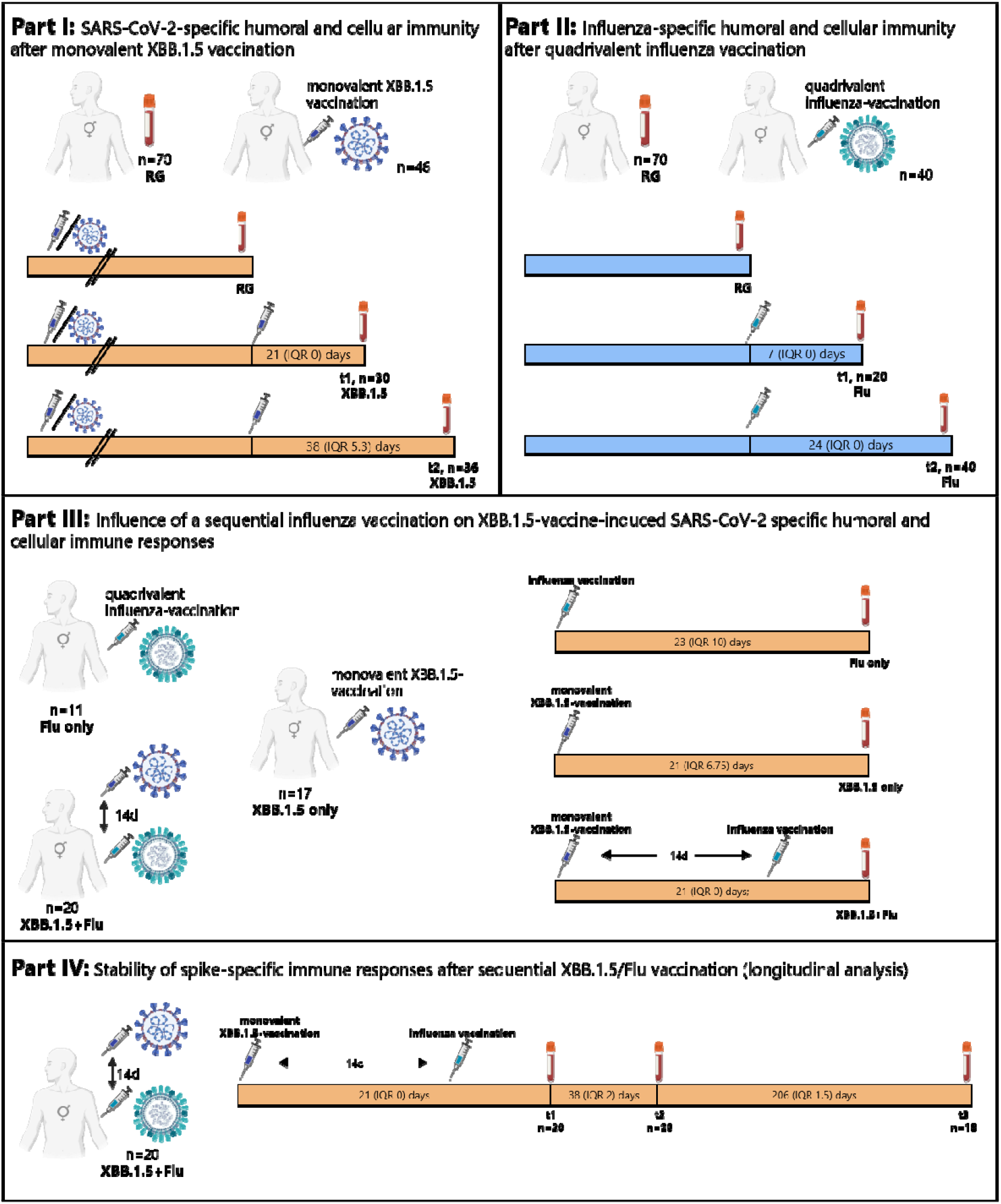
Schematic overview of the four parts of the study. Dialysis patients were recruited in a cross-sectional study for analysis of baseline immunity before the vaccine season (n=70, reference group; RG). A total of 57 patients were tested after having received either the monovalent XBB.1.5 vaccination alone (n=17), influenza vaccination alone (n=11), or sequential administration of both vaccines (n=29, with the influenza vaccine administered fourteen days after XBB.1.5 vaccination). **Part I:** SARS-CoV-2-specific humoral and cellular immune responses were characterized in 46 patients after monovalent XBB.1.5-vaccination irrespective of influenza vaccination (30 patients after a median of 21 (IQR 0) days (“XBB.1.5 t1”), 36 patients after a median of 38 (IQR 5.3) days (“XBB.1.5 t2”)) and compared with the data from the reference group (n=70). **Part II:** Influenza-specific humoral and cellular immune responses were characterized in 40 patients after influenza vaccination irrespective of XBB.1.5 vaccination (20 patients after a median of 7 (IQR 0) days (“Flu t1”), 40 patients after a median of 24 (IQR 0) days (“Flu t2”) and compared with the data from the reference group (n=70)). **Part III:** SARS-CoV-2-specific immune responses were compared in dialysis patients after monovalent XBB.1.5 vaccination only (n=17; “XBB.1.5 only”), influenza vaccination only (n=11; “Flu only”), or sequential administration of XBB.1.5 followed by influenza vaccination (n=20; “XBB.1.5+Flu”). Testing was performed approximately three weeks after the XBB.1.5 vaccination as indicated in the figure. **Part IV:** The stability of the SARS-CoV-2-specific immune response in 20 dialysis patients after sequential administration of XBB.1.5 followed by influenza vaccination was investigated over a period of 6 months with time points indicated in the figure. Two patients were lost to follow-up (1 died, 1 was transplanted).

**Supplementary figure S2:**
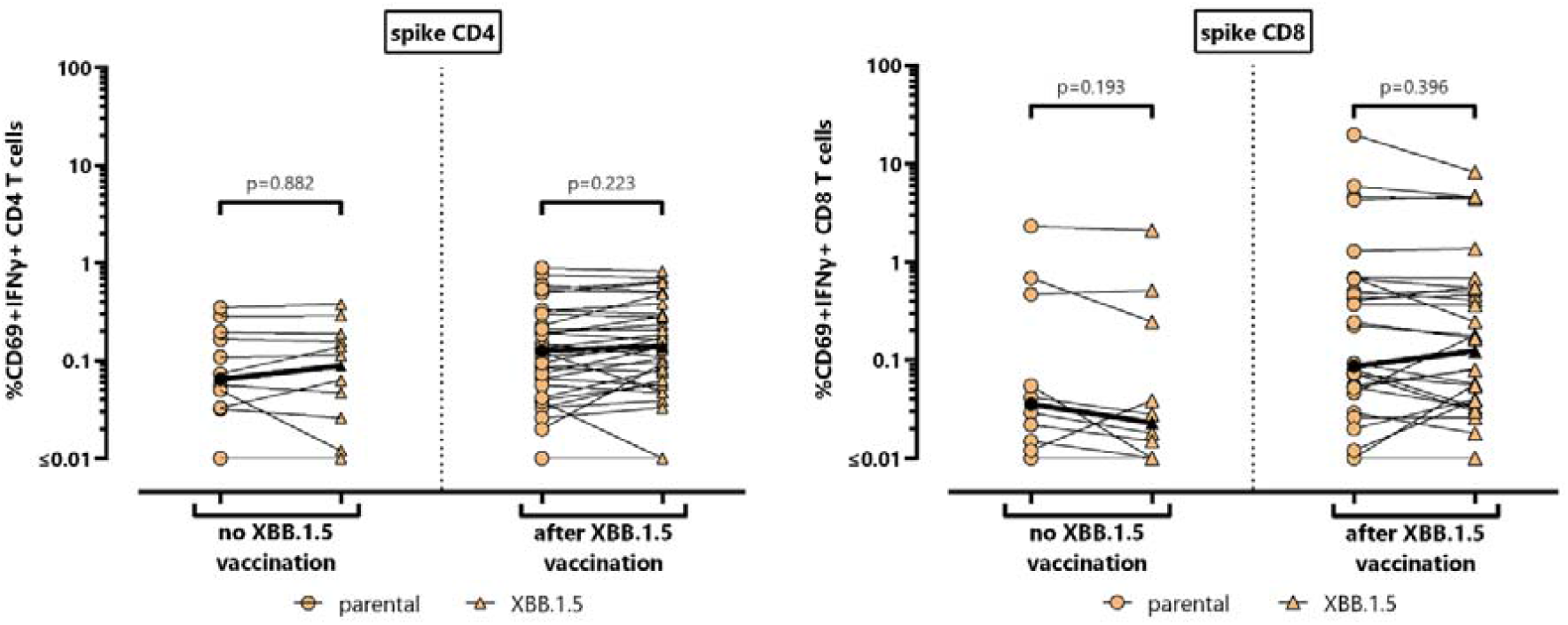
Comparison of CD4 and CD8 T-cell reactivity towards spike from the parental strain and the Omicron variant XBB.1.5. Levels of CD4 and CD8 T cells reactive towards parental spike (circle) and spike from the Omicron subvariant XBB.1.5 (triangle) were compared in subgroups of dialysis patients without (n=12) and after monovalent XBB.1.5 vaccination (n=34). Median levels are indicated. Differences among paired datasets were calculated by Wilcoxon signed rank test. Abbreviations: IFN, interferon

