## Supplementary figures for "SARS-CoV-2-specific immunity after XBB.1.5 vaccination is not influenced by subsequent influenza vaccination in dialysis patients"

The supplement contains 2 supplementary figures S1 and S2.

### Supplementary figures

#### Supplementary Figure S1

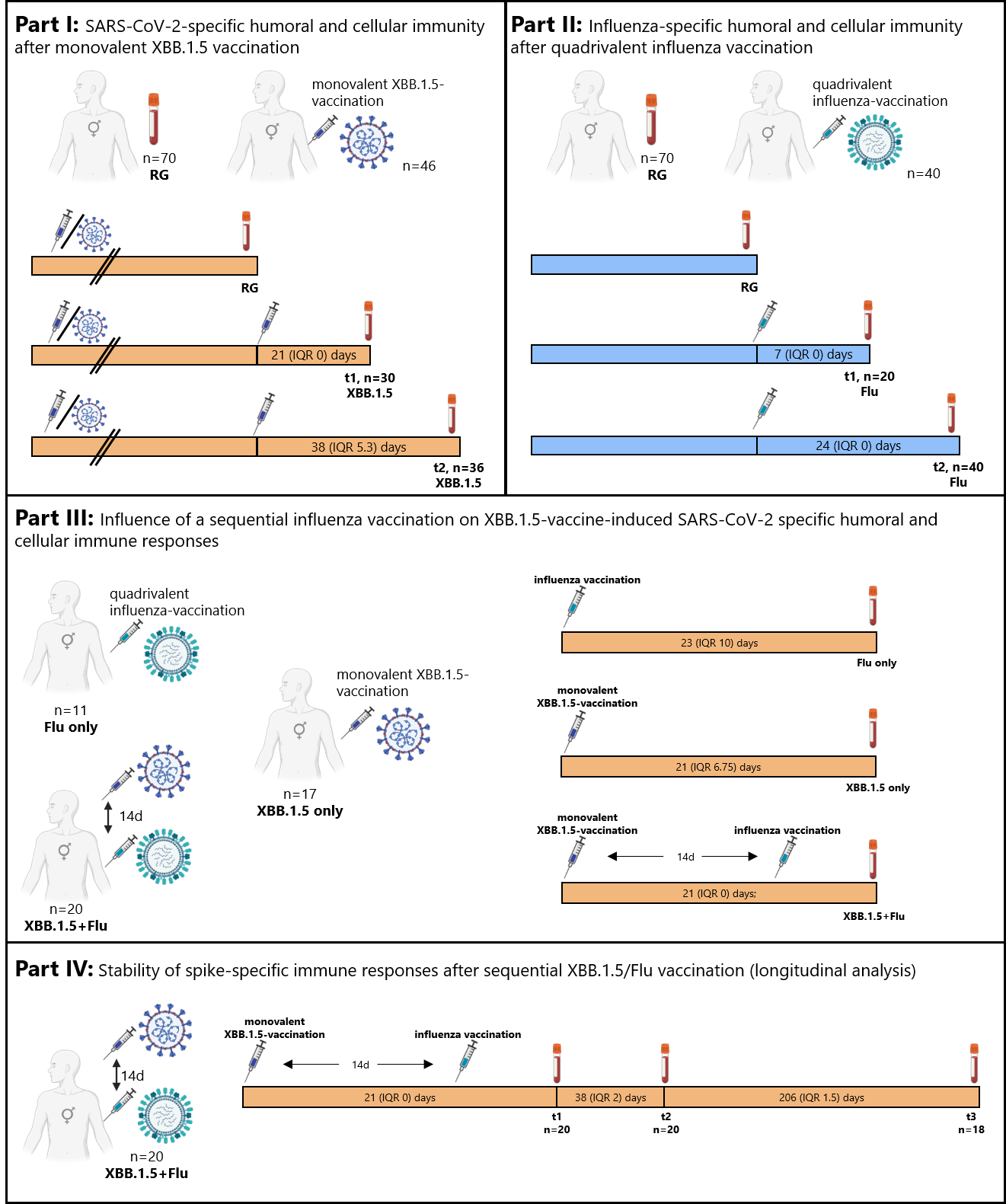

**Supplementary figure S1. Schematic overview of the four parts of the study.** Dialysis patients were recruited in a cross-sectional study for analysis of baseline immunity before the vaccine season (n=70, reference group; RG). A total of 57 patients were tested after having received either the monovalent XBB.1.5 vaccination alone (n=17), influenza vaccination alone (n=11), or sequential administration of both vaccines (n=29, with the influenza vaccine administered fourteen days after XBB.1.5 vaccination). **Part I:** SARS-CoV-2-specific humoral and cellular immune responses were characterized in 46 patients after monovalent XBB.1.5-vaccination irrespective of influenza vaccination (30 patients after a median of 21 (IQR 0) days (“XBB.1.5 t1”), 36 patients after a median of 38 (IQR 5.3) days (“XBB.1.5 t2”)) and compared with the data from the reference group (n=70). **Part II:** Influenza-specific humoral and cellular immune responses were characterized in 40 patients after influenza vaccination irrespective of XBB.1.5 vaccination (20 patients after a median of 7 (IQR 0) days (“Flu t1”), 40 patients after a median of 24 (IQR 0) days (“Flu t2”) and compared with the data from the reference group (n=70)). **Part III:** SARS-CoV-2-specific immune responses were compared in dialysis patients after monovalent XBB.1.5 vaccination only (n=17; “XBB.1.5 only”), influenza vaccination only (n=11; “Flu only”), or sequential administration of XBB.1.5 followed by influenza vaccination (n=20; “XBB.1.5+Flu”). Testing was performed approximately three weeks after the XBB.1.5 vaccination as indicated in the figure. **Part IV:** The stability of the SARS-CoV-2-specific immune response in 20 dialysis patients after sequential administration of XBB.1.5 followed by influenza vaccination was investigated over a period of 6 months with time points indicated in the figure. Two patients were lost to follow-up (1 died, 1 was transplanted).

#### Supplementary figure S2

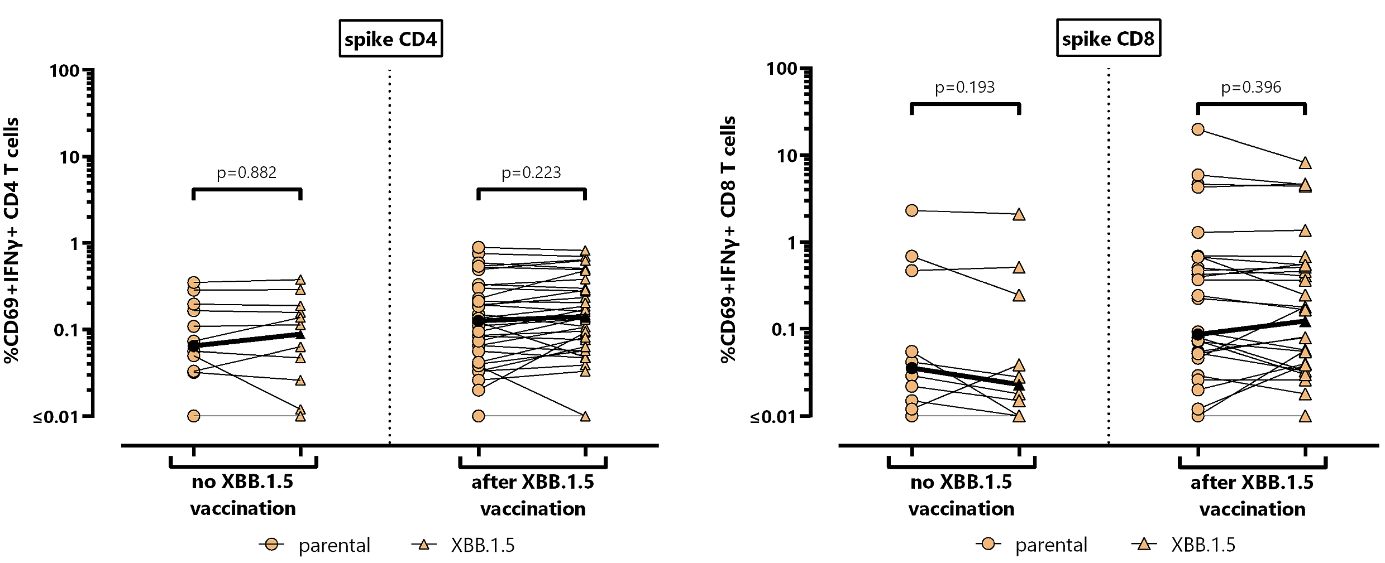

**Supplementary figure S2: Comparison of CD4 and CD8 T-cell reactivity towards spike from the parental strain and the Omicron variant XBB.1.5.** Levels of CD4 and CD8 T cells reactive towards parental spike (circle) and spike from the Omicron subvariant XBB.1.5 (triangle) were compared in subgroups of dialysis patients without (n=12) and after monovalent XBB.1.5 vaccination (n=34). Median levels are indicated. Differences among paired datasets were calculated by Wilcoxon signed rank test. Abbreviations: IFN, interferon
